# Comparison of student perception and exam validity, reliability and items difficulty: cross-sectional study

**DOI:** 10.1101/19002535

**Authors:** Assad Ali Rezigalla, Ali Mohammed Elhassan Eleragi, Masoud Ishag Elkhalifa, Ammar M A Mohammed

## Abstract

**Introduction:** Student perception of an exam is a reflection of their feelings towards the exam items, while item analysis is a statistical analysis of students’ responses to exam items. The study was formulated to compare the student’s perception of the results of item analysis.

**Material and methods:** Type of the study is cross-sectional. The study was conducted in the college of medicine, in the duration from January to April 2019. The study uses a structured questionnaire and standardized item analysis of students’ exam. Participants are students registered for semester two level year (2018-2019). Exclusion criteria included all students who refused to participate in the study or do not fill the questionnaire.

**Result:** The response rate of the questionnaire was 88.9% (40/45). Students considered the exam as easy (70.4%). The average difficulty index of the exam is acceptable. KR-20 of the exam was 0.906. A significant correlation was reported between student perceptions towards exam difficulty and standard exam difficulty.

**Discussion:** Student perceptions support the evidances of exam vlaitdity. Students can estimate exam difficulty.

## Introduction

Students’ perception of an exam is a reflection of their feelings towards the exam items, while item analysis is a statistical analysis of students’ responses on exam items. Whether students’ perception or item analysis is agreed or not, they represent different views about exam items.

Students’ perception is widely used and recommended in medical education. Data generated from students’ perception can provide information about faculty, the achievement of educational objectives, and instructional methods (1, 2). Also, it is considered as a reliable and valid indicator of effective teaching (3).

Construct validity denoted as “a unitary concept, requiring multiple lines of evidence, to support the appropriateness, meaningfulness of the specific inferences made from test scores”(4). Thus, the assessment is considered valid if measures what is intended to measure and reflects the educational contents (5, 6). The mismatch between the level of the cognitive process in the assessment and the educational task can affect exam validity and reliability (5, 7). The mismatch can appear in the form of too many easy or difficult items.

Item analysis is a mathematical analysis of students’ responses on an exam to evaluate items quality and consequently improving the assessment. This can be done either by refining the defected or deletion of poorly constructed from the questions bank (8-10). Quality of items is evaluated through a variety of item analysis parameters, which include Difficulty Index (DIF) and the index of the internal consistency (KR-20). DIF is defined as the percentage of the examinees who answered the item correctly. It ranges from 0% to 100%; with a higher value indicating an easy item (11).

Commonly the internal consistency is measured through Cronbach’s α. (Coefficient alpha) (8, 12, 13). Coefficient alpha is identical to Kuder–Richardson formula 20 (KR-20) when each item has a single answer (MCQs Type A)(8, 14, 15). There are different ranges and interpretations of item analysis parameters and internal consistency published in the literature (12, 15-20) (Table 1).

**Table 1:** Classification of exam items according to difficulty index.

| Parameters | UBCOM |  | (16) |  | (19) |  |
| --- | --- | --- | --- | --- | --- | --- |
|  | Interpretation | % | Interpretation | % | Interpretation | % |
| DI | Easy (<80) | 35 | Easy (>70) | 52.5 | Easy (>80) | 35 |
|  | Moderate (25-80) | 62.5 | Acceptable (30-70) | 42.5 | Acceptable (30-80) | 60 |
|  | Difficult (0-25) | 2.5 | Difficult (<30) | 5 | Difficult (30<) | 5 |

DI, Difficulty index.

The college of Medicine, adopts SPICE curriculum. Problem-based learning is the principal educational strategy as well as an instructional method. The program offers an MB, BS after successful completion of twelve semesters (six years)(21, 22).

This study was conducted to compare student’s perception towards exam validity, reliability, and difficulty.

## Materials and Methods

### Study design

The study is a cross-sectional study. It was conducted at college of medicine, during the period from January to April 2019.

### Materials

The study used a questionnaire to evaluate student’s perception towards the exam items and standard item analysis of the exam.

The questionnaire was developed to gain a deep understanding of students’ perceptions towards the exam generally and exam items individually. It was developed by the authors and consultation of medical educationalist and satiation and consisted of two parts. Part one investigates the levels of items difficulty (easy, moderate, and difficult) and whether the specific learning outcome (SLO) from which the items were constructed, were covered or not. The second part encompasses the number of items, their mode of covering the course contents and the ability of exam to assess students. The questionnaire was tested through a pilot study. Data generated from the pilot study were not included in the study.

The exam used in this study was the exam of principles of human diseases course. It conducted in semester two-second year (n=45). The course is integrated and multi-disciplinary. The course exam was developed by the course committee using course blueprint and then approved by the assessment committee. It was formed of MCQs type A. The number of exam items (n=80) was adjusted according to the course blueprint and the tested domains (23). Each item is composed of stem and four options, three distractors, and a single best answer. The correct answer is awarded one mark and no marks for blank or wrong selection.

The exam was marked (DataLink 1200 - Apperson) and double checked. Standard item analysis was obtained and processed for the study.

### Participants

All students registered for the course of principles of human diseases (2018-2019) were included in the study. Exclusion criteria included students who refused to participate in the study or do not fill the questionnaire. Students filled the questionnaire immediately after completing the exam without identification.

### Ethical consideration

The study was approved by research and ethics committee college of medicine. All students accepted to participate in the study filled a written consent.

### Statistical analyses

The data obtained from both of the questionnaires and the standard item analyze analyzed by using SPSS V20 (Armonk, NY: IBM Corp, USA). Descriptive statistics and Pearson correlation coefficient were applied to measure the significance of difference and correlation among different variables. Level of significance was fixed at 95%, and any *P* < 0.05 was considered to be significant.

## Results

### Student’s perceptions

The response rate was 88.9% (40/45). The average students’ perceptions of exam items were easy (70.4%), moderate difficulty (18.5%) and difficult (11.1%). All most all of the students (92.3%) reported that the exam items were covered as an objective during the course. For 57% of the students, the exam items cover the entire course content. 50% of the students reported that exam items were not concentrated on certain topic, while, 43% reported that exam items were concentrated, where 7% of the students were not sure. For 70% of the students, the number of test item was adequate to assess them. On the other hand, 43% of the students considered the test, in general, is adequate to evaluated students and 50% reported not adequate and the remaining (7%) were not sure (Figure 1).

**Figure 1:**
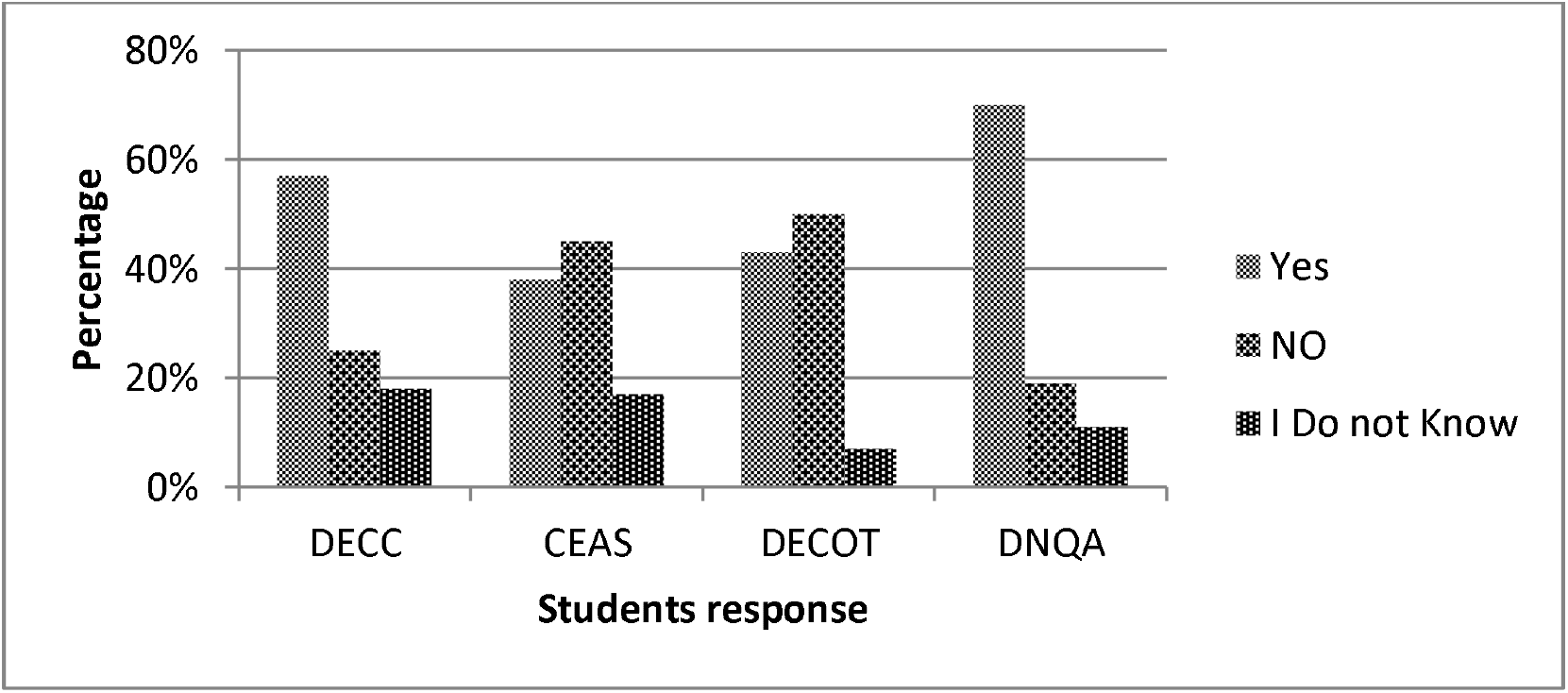
Students perceptions towards exam. DECC, Does the exam cover the entire course; CEAS, Can the exam assess students; DECOT, Does the exam concentrate on certain topics; DNQA, Does the number of question is adequate to assess students.

The correlation between average student perceptions of items difficulty the difficulty index of exam items were significant. A moderate positive correlation was reported between easy perception and DIF (r=0.7033, p=.00001), which means there is a tendency for high DIF go with high easy perception (and vice versa). Moderate negative correlation reported between moderate (r=-0.2969, p= .008082) and difficult (r= −0.6094, p= .00001) student perceptions and DIF (Graph 1).

A significant moderate positive correlation (r=.615, p=.00001) was reported between DIF and items from covered SLOs (Graph 2).

### Item analysis

The total number of the analyzed items was 80. The average score of the class was 55.5 (69.38%). Class median was 56.0 (70.0%). KR-20 was 0.906. Students pass rate in the exam was 32.5% (Pass mark=60). The average DIF of the exam was 69.4 (±21.86). Items were classified according to their difficulty in difficult, moderate (acceptable) and easy (Table 1).

## Discussion

About 92.3% of the students reported that exam items were from course SLOs. Half of the students reported that the exam covers the course contents. Furthermore, 70% of the students believe that the exam can assess students.

Approving of exam validity depends on the evidence that can support validity. (5, 6, 18). The methodology of exam construction (23) expert staff member who involved in the course teaching support the exam validity. Student perceptions in regards to the exam in general, support that the exam measures what is intended to measure and reflect the educational contents. These findings are by previous work of Carmines and Zeller (18, 24)

KR-20 of the exam was 0.906. Classification of exam items shows that most of the items were within the acceptable range of difficulty (62.5%), and only 37.5 % of items were not. It has been reported that the presence of too many easy or difficult items can affect both exam validity and reliability(5, 7). According to some authors (5, 12, 17, 20, 25) values of KR-20 such as 0.8 or above is ideal and demonstrate excellent reliability of exam and the target goal in clinical practice.

The average DIF of exam according to the standard item analysis was 69.4. The average student’s perception of exam difficulty is easy (70.5%) and has a significant positive correlation (p=0.001) with DIF. The average exam difficulty is considered as good and acceptable according to College assessment policy and literature (16, 17, 19). In any exam or test, the average difficulty of items should be adjusted according to the required competencies and students level, and these two points are the areas of concerns for item constructors or assessment composers (5). The current findings of student’s perception towards exam difficulty suggested that they underestimated the exam difficulty. Students commonly underestimate their performance rather than the exam difficulty(5, 26, 27). Van de Watering reported that student’s perception toward exam difficulty is differing according to the student performance in the exam(5). Students with higher scores underestimate their performance.

Meanwhile, those with lower scores have more accurate estimations. According to the exam result, the upper students represent 72.5%. However, the class mean and average are relatively similar (55.5 and 56.0, respectively). The result suggested a good student’s performance. These findings support the work of Van de Watering(5).

The limitation of the study includes the fewer number of students and application on one course. The strength of the study, the test is considered valid and reliable through several pieces of evidence.

## Conclusion

Student perceptions can support exam validity and reliability. Students can estimate test difficulty, although they were reported to underestimate their performance.

## Data Availability

Data will be available upon request to the corresponding author.

## Acknowledgment

The author acknowledges the students who participated in the study. Great appreciation was to Dr. Elwathiq Khalid, Dr. K. Salih, Dr. E. Miskeen, Dr. A.MS. Eleragi, Dr. I. Jack, Prof. Masoud Ishag (College of Medicine, University of Bisha) and the appreciation extend to Dr. M. Elhassan (College of Medicine, Qassim University, Saudi Arabia) and my colleagues. Great thanks to Mr. MK. Abid (College of Medicine, King Khalid University, Abha, Saudi Arabia) for the statistical analysis and helpful comments. Special thanks and appreciation to College Dean and Administration of the College of Medicine, University of Bisha (Bisha, Saudi Arabia) for help and allowing the use of facilities.

---

**Graph 1:**
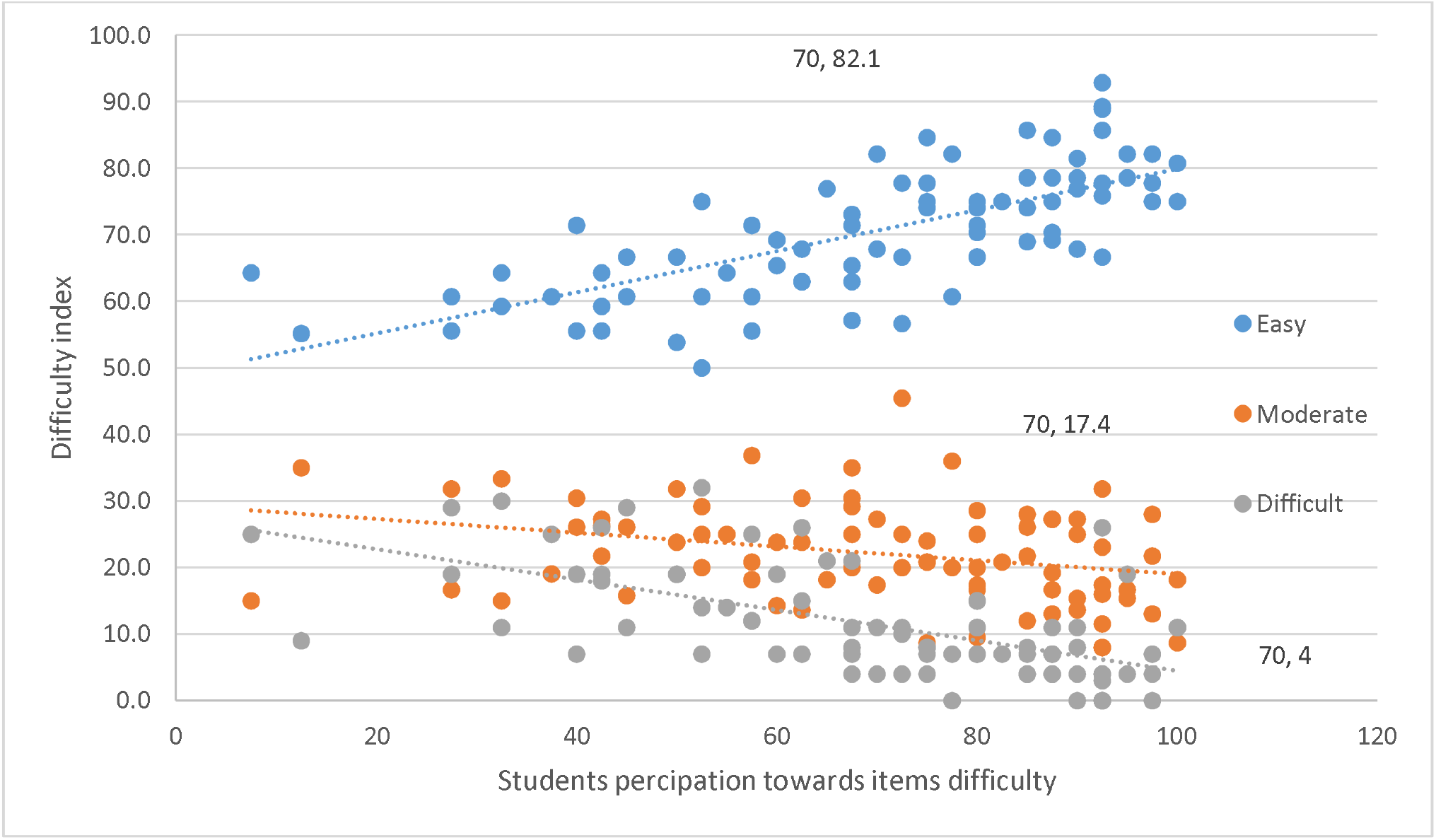
Correlation of student’s perceptions towards item difficulty and the standard difficulty index of items.

**Graph 2:**
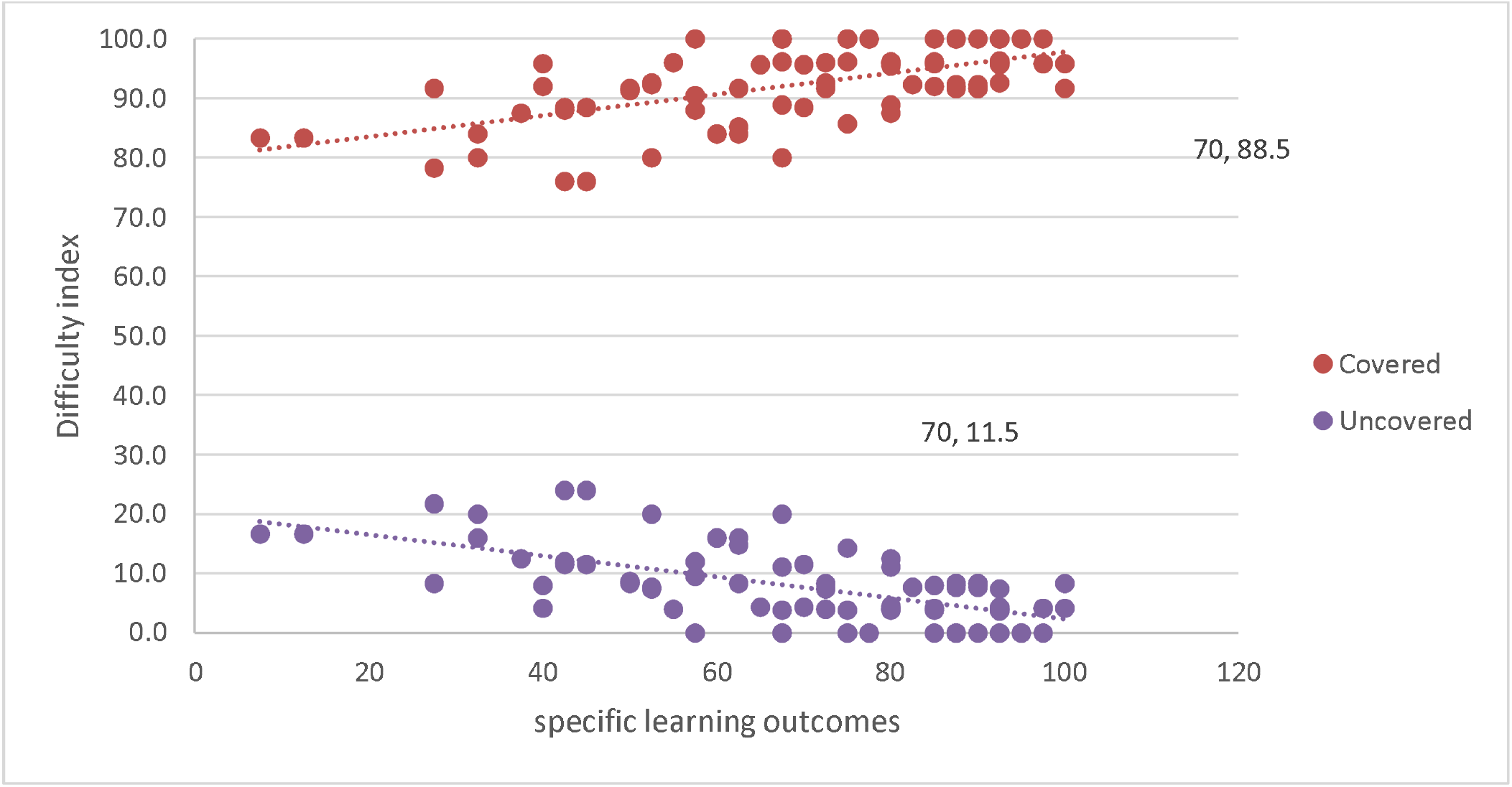
The Correlation between difficulty index of items and the specific learning outcome.

